# Strengthening Patient Safety in a Cambodian Hospital Through Structured Weekly Reporting: A Low-Cost Quality Improvement Initiative

**DOI:** 10.1101/2025.10.26.25338511

**Authors:** Suren Kanayan

**Author notes:** **Ethics Statement** This project met institutional criteria for quality improvement and did not require formal ethics approval. All data aggregated and anonymized. **Funding Statement** No external funding was used.

## Abstract

Persistent patient safety issues in low-resource hospitals are often underreported, delaying corrective actions and increasing preventable harm. This project aimed to implement a low-cost, structured weekly reporting system to improve patient safety processes and outcomes across departments at a tertiary hospital in Phnom Penh, Cambodia.

The structured weekly reporting system was designed and implemented by the author, with departmental staff providing operational input. A one-page Departmental Wise Checklist was introduced in the Emergency Room/Intensive Care Unit (ER/ICU), Obstetrics and Gynecology (OB/GYN), Outpatient Department (OPD), and Pediatrics. Weekly reviews, feedback loops, and departmental champions supported consistent reporting and follow-through.

After three months, process measures improved substantially: formally adopted SOPs increased from 0 to 10, BLS-trained ER staff rose from 38% to 98%, incidents of expired blood products fell from two per month to none, and reporting compliance averaged 92%. Patient-level outcomes also improved: monthly cases of neonatal hypothermia decreased from six to two, and ER adverse events dropped from four to one.

This initiative demonstrates that structured weekly reporting, combined with iterative feedback and departmental engagement, can strengthen patient safety and accountability in low-resource settings. Sustaining improvements requires embedding the process into quality assurance activities, staff training, and continued leadership oversight.

**Key Messages:**

- In low-resource hospitals, patient safety systems often lack structured reporting and accountability mechanisms.
- A low-cost weekly reporting system refined through iterative PDSA cycles can improve engagement, compliance, and safety issue resolution across multiple departments.
- This model provides a replicable framework for hospitals in resource-limited settings to strengthen governance and foster a culture of safety.

## Introduction

Persistent patient safety challenges observed in a tertiary hospital in Phnom Penh, Cambodia. These included expired blood units in the emergency department, inconsistent clinical documentation, and communication gaps during shift handovers. At baseline, safety reporting was informal and unstructured, resulting in limited visibility for leadership into systemic vulnerabilities.

The hospital serves a diverse urban patient population, with multiple high-acuity departments including Emergency/ICU, Obstetrics and Gynecology, Pediatrics, and Outpatient services. Staff numbers are constrained, and workflows are often stretched due to patient volume and limited resources.

The focus of this project was to implement a low-cost, sustainable mechanism for frontline staff to report patient safety concerns, ensure timely escalation, and track follow-through. The SMART aim was to achieve >90 % structured safety reporting compliance and a ≥50 % reduction in key adverse events (expired blood products, neonatal hypothermia) within 12 weeks. Achieving this would provide leadership with actionable insights, strengthen accountability, and improve patient safety culture.

## Background

Patient safety is a fundamental component of healthcare quality, yet in low- and middle-income countries (LMICs) formal reporting mechanisms for safety incidents are often absent or inconsistently applied. [1,2] This results in underreporting of adverse events, delayed corrective actions, and ongoing preventable harm to patients. Evidence from LMIC hospitals indicates that recurring safety risks—such as expired blood products, inconsistent documentation, and communication breakdowns during handovers—frequently remain undetected without a structured reporting system. [2,3]

Previous studies have demonstrated that structured reporting interventions can improve the recognition and management of safety hazards. For example, Pronovost et al. [6] showed that implementing standardized reporting and feedback in intensive care settings reduced catheter-related bloodstream infections and improved adherence to care protocols. Herzer et al. [7] reported sustained improvement in patient safety metrics through multidisciplinary reporting teams combined with “Good Catch” awards, highlighting the importance of feedback and staff engagement. Evidence also suggests that combining structured reporting with departmental ownership, iterative review, and feedback loops is essential for sustainable improvement. [4,5]

Attempts to improve patient safety in resource-limited settings face specific challenges. Kruk et al. [3] and Pichumani et al. [2] emphasize that incomplete reporting, lack of ownership, and absent feedback loops diminish the impact of interventions. Staff may lack time, experience, or incentives to submit reports, and leadership may have limited capacity to monitor systemic vulnerabilities. Without mechanisms to capture and act on safety data, hospitals risk persistent process failures, including delays in addressing expired medications or blood products, inconsistent implementation of SOPs, and preventable adverse events.

Few published reports from Southeast Asia describe sustainable, low-cost mechanisms for real-time safety reporting integrated into hospital governance structures. In the context of a tertiary hospital in Phnom Penh, Cambodia, these challenges were evident: safety incidents were recorded ad hoc, and there was no structured mechanism to escalate or resolve issues. This gap limited leadership oversight and hindered systematic process improvement. Structured weekly reporting using a simple, one-page departmental checklist has been proposed as a low-cost, feasible intervention to capture actionable safety information, enhance accountability, and provide a platform for iterative improvement. [6,7] By combining standardized data collection, departmental champions, and governance oversight, hospitals can improve team communication, increase adherence to SOPs, and reduce preventable adverse events. Evidence indicates that these strategies, when implemented with ongoing feedback, can yield measurable improvements in both process and patient-level outcomes even in low-resource hospital contexts. [4,5]

## Measurement

To monitor both process and patient-level outcomes, a one-page Departmental Wise Checklist was collaboratively developed with departmental leads. The checklist captured key domains:

- Patient care issues (e.g., expired blood products, near-miss events)
- Staffing constraints and coverage gaps
- Documentation errors or omissions
- Equipment, supply, and logistical shortages

Baseline data were collected retrospectively for the six months preceding the intervention from multiple sources, including blood bank logs, departmental records, training registries, neonatal unit logs, and incident reports. At baseline, no structured reporting system existed, and leadership had limited visibility into recurring safety issues. Key baseline metrics included:

- Expired blood units (ER): 2 per month
- Formal SOPs: none implemented for intrapartum monitoring across departments
- BLS-trained ER staff: 38%
- Neonatal hypothermia (OB/GYN): 6 cases per month
- ER adverse events: 4 per month
- Reporting compliance: negligible, as no structured system existed

Rationale for measure selection: Metrics were chosen to reflect both process adherence (SOP adoption, BLS training, reporting compliance) and patient safety outcomes (expired blood products, neonatal hypothermia, ER adverse events). Operational definitions were as follows:

- Expired blood units: any blood product past expiry discovered in ER inventory
- SOP adoption: number of departments with formal, documented SOPs actively implemented
- BLS-trained staff: percentage of ER personnel with up-to-date certification
- Neonatal hypothermia: number of newborns with axillary temperature <36.5°C within 24 hours of?birth
- ER adverse events: incidents reported through the hospital incident reporting system
- Reporting compliance: proportion of weekly checklists submitted on time by each department

During the 12-week intervention period, data were collected weekly. The project team cross-checked checklist submissions with departmental records and incident logs to ensure completeness and accuracy. Descriptive statistics and run charts were used to display trends over time. Due to small numbers, formal statistical process control methods were not applied.

Baseline measurements highlighted substantial gaps in both process and patient-level outcomes, confirming the need for a systematic intervention to improve patient safety and accountability.

## Design

The quality improvement initiative was led by author, who designed, implemented, and monitored the structured weekly reporting system. Departmental participation came from head nurses and leads of ER/ICU, Obstetrics & Gynecology, Pediatrics & Neonatology, and OPD, providing input for checklist completion and reporting, under oversight of the Hospital Quality Improvement Committee. The goal was to create a standardized, low-cost mechanism for frontline staff to report patient safety issues, trigger timely corrective actions, and monitor follow-through. Simplicity and clarity were prioritized, as complex tools could hinder adoption in a busy, resource-constrained environment.

A one-page Departmental Wise List (a structured checklist for each department) was developed, combining tick-box items with optional short notes to capture urgent, ongoing, or deferrable concerns across patient care, documentation, staffing, and equipment. Weekly submission to the hospital Quality Improvement (QI) Committee was standardized to ensure governance oversight and accountability.

To assess feasibility and usability, a two-week pilot was conducted in the Emergency Room/Intensive Care Unit (ER/ICU). Feedback from frontline staff was used to refine wording, reduce reporting burden, and optimize submission timing.

Sustainability was ensured by embedding the Wise List into routine departmental workflows. Departmental champions were appointed to maintain momentum, provide coaching, and escalate unresolved issues. Oversight by the hospital QI Committee provided ethical guidance and operational accountability, reinforcing a culture of safety and continuous improvement. Process mapping illustrated the flow of information from Wise List completion to committee review, showing how the intervention streamlined reporting, improved interdepartmental communication, and accelerated corrective actions.

The intervention required approximately 1 staff hour per department per week for data entry and review, with all documentation completed electronically in line with the hospital’s green initiative. Weekly QI committee meetings lasted about 30 minutes, representing minimal additional cost.

## Strategy

The overarching aim of the project was to implement a structured, low-cost weekly reporting system to improve patient safety and accountability across multiple departments, including the ER/ICU, OB/GYN, Pediatrics, and OPD. To achieve this, a series of Plan-Do-Study-Act (PDSA) cycles employed, iteratively refining the intervention based on real-world feedback.

PDSA Cycle 1: The Departmental Wise List drafted, and department leads were oriented to its purpose and use. The Wise List piloted in the ER/ICU for two weeks. Observations and staff feedback revealed ambiguous items, barriers to timely submission, and concerns about workload, which limited engagement.

PDSA Cycle 2: The Wise List was simplified. Tick boxes emphasized, free-text responses minimized, and brief coaching sessions clarified its intent and use. Weekly submission compliance improved, though early submissions remained inconsistent, averaging 75% during the pilot.

PDSA Cycle 3: Implementation expanded to OB/GYN, Pediatrics, and OPD. Weekly QI committee meetings provided oversight, classified reported issues, assigned ownership, and tracked resolution. Escalation fatigue emerged as a challenge, as recurrent issues persisted without resolution. To address this, departmental champions appointed to ensure follow-up and accountability. Reported items were categorized by urgency to reduce data overload and focus attention on high-priority patient safety concerns.

Continuous review of performance data, frontline feedback, and iterative refinements ensured that actionable safety concerns were systematically addressed. Overall, this strategy strengthened engagement, enhanced reporting compliance, and fostered a culture of accountability. These experiences provide practical guidance for other low-resource hospitals seeking to implement similar quality improvement interventions.

## Results

Process and Patient-Level Outcomes After Three Months

Over the 12-week intervention period, the implementation of the weekly reporting system led to substantial improvements in both process measures and patient-level outcomes. At baseline, no formal SOPs were in place, only 38% of ER staff were trained in Basic Life Support (BLS), and reporting compliance was negligible.

By week 12, formal SOP adoption increased from 0 to 10 across the ER, OB/GYN, and other departments (Figure 1), reflecting structured process improvements and better standardization of care. BLS training coverage in the ER improved markedly, rising from 38% to 98% after retraining 24 staff members (Figure 1), demonstrating enhanced readiness for emergency situations. Departmental compliance with weekly checklist submissions averaged 92% over the 12-week period (Figure 1), indicating successful integration of structured reporting into routine workflows. Interdisciplinary huddles were established in all departments, providing forums for reviewing safety issues, assigning responsibility, and accelerating problem resolution (Table 1). For instance, OB/GYN developed and implemented an intrapartum monitoring SOP within one month, and laboratory turnaround protocols were optimized (Table 1).

**Table 1.**
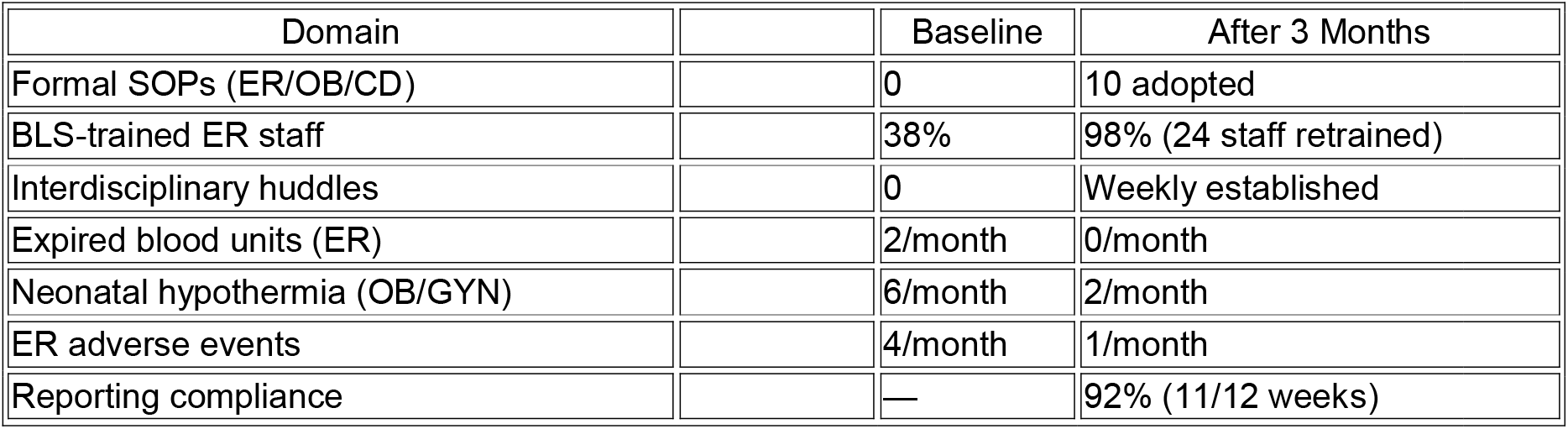
Baseline and 3-Month Outcomes.

| Domain |  | Baseline | After 3 Months |
| --- | --- | --- | --- |
| Formal SOPs (ER/OB/CD) |  | 0 | 10 adopted |
| BLS-trained ER staff |  | 38% | 98% (24 staff retrained) |
| Interdisciplinary huddles |  | 0 | Weekly established |
| Expired blood units (ER) |  | 2/month | 0/month |
| Neonatal hypothermia (OB/GYN) |  | 6/month | 2/month |
| ER adverse events |  | 4/month | 1/month |
| Reporting compliance |  | — | 92% (11/12 weeks) |

**Figure 1.**
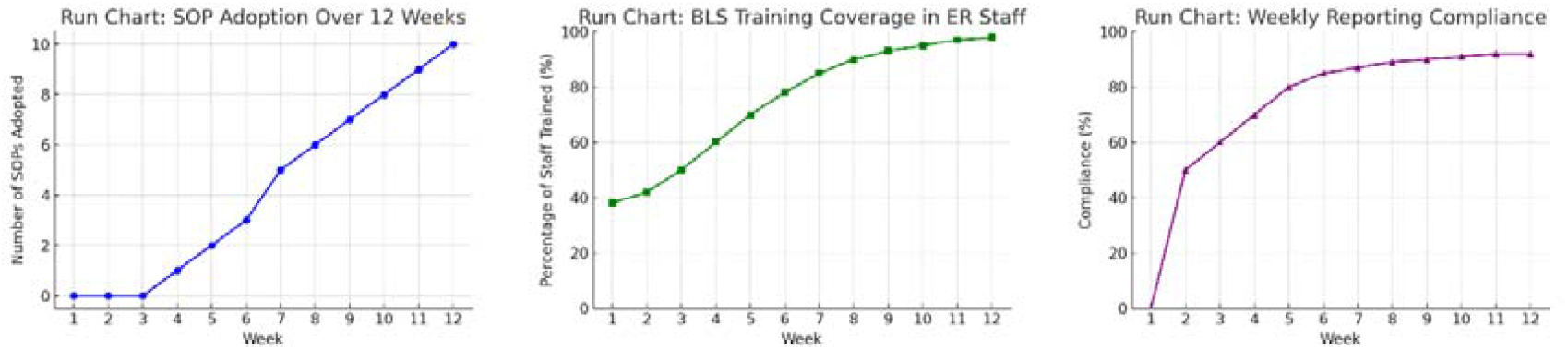
Illustrates trends in process measures over the 12-week intervention period, including SOP adoption, BLS training coverage, and weekly reporting compliance.

Patient-level outcomes also improved over the 12-week period (Figure 2). Expired blood units in the ER decreased from 2 per month to 0, neonatal hypothermia cases declined from 6 to 2 per month, and ER adverse events fell from 4 to 1 per month (Figure 2), demonstrating measurable improvements in patient safety following the intervention.

**Figure 2.**
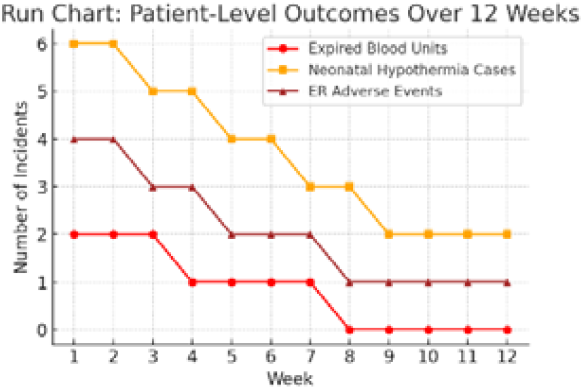
Displays patient-level outcomes over the same period, including expired blood units in the ER, neonatal hypothermia cases, and ER adverse events. These trends show clear improvement compared with baseline measurements.

These results indicate that a low-cost, structured weekly reporting system—combined with iterative feedback, departmental champions, and oversight by the hospital QI committee— can produce measurable improvements in both process and patient safety outcomes in low-resource hospital settings. Integration of quantitative metrics, visual run charts, and narrative documentation of challenges reinforced accountability, encouraged continuous improvement, and facilitated timely resolution of safety issues. Interdisciplinary huddles played a key role in this success, fostering better communication and rapid problem-solving across departments. The structured SOP implementation, along with optimized lab and clinical protocols, ensured that improvements were sustained throughout the intervention period. Frontline staff reported improved communication and faster resolution of issues during weekly huddles.

## Lessons

This project demonstrated that structured weekly reporting can foster transparency, accountability, and measurable improvements even in low-resource hospital settings. Narrative documentation of challenges, iterative adaptations, and clear assignment of ownership were as important as quantitative metrics in ensuring the intervention’s effectiveness. Feedback loops, supported by departmental champions, were critical for timely follow-through and sustainability, while governance oversight by the hospital QI Committee maintained ethical rigor and operational accountability.

Several lessons emerged. Engaging frontline staff in checklist design increased buy-in and facilitated integration into routine workflows. Iterative refinement through pilot testing and PDSA cycles allowed identification and resolution of practical barriers, such as ambiguous items, submission delays, and workload concerns. The appointment of departmental champions to oversee checklist completion and follow-up ensured that improvements were sustained beyond the initial implementation phase.

## Limitations

Limitations of the project include its single-site, short-term design, which may limit generalisability to other hospitals with different staffing levels, patient populations, or resources. Patient-level outcomes were restricted to selected measurable events, such as neonatal hypothermia and expired blood units, and broader morbidity or mortality impacts were not assessed. Long-term sustainability depends on continued embedding of structured reporting into routine quality assurance processes, consistent leadership oversight, and ongoing staff training. External factors, such as staff turnover and variation in departmental workloads, may have influenced results independently of the intervention.

Finally, this project demonstrates that low-cost, process-oriented interventions can yield meaningful safety improvements in LMIC hospitals. Key factors contributing to success included active frontline staff engagement, iterative refinement through PDSA cycles, clear assignment of departmental responsibility, and integration into routine workflows. While these principles are transferable to other low-resource settings, adaptation to the local context is essential. Regional evidence supports similar approaches in Southeast Asia, showing that structured reporting combined with feedback and departmental ownership improves patient safety and care quality. [2,6,7,8,9] Future cycles will include staff satisfaction as a balancing measure to ensure sustainability without added burden.

## Conclusion

This project demonstrates that implementing structured weekly reporting, supported by narrative documentation of planning, challenges, and iterative adaptations, can significantly strengthen patient safety and accountability in a low-resource hospital setting. Our SMART aim—to introduce a low-cost, sustainable reporting mechanism that improves process compliance and patient-level outcomes—was largely achieved: formal SOP adoption increased from 0 to 10, BLS training coverage rose from 38% to 98%, expired blood units were eliminated, and neonatal hypothermia and ER adverse events declined.

The measures selected were appropriate to capture both process and patient-level outcomes, and the use of departmental champions, iterative PDSA cycles, and weekly QI oversight ensured follow-through and sustained engagement. The intervention was low-cost, requiring minimal additional resources, and resulted in indirect financial savings by reducing waste from expired blood products and improving operational efficiency.

Sustainability is supported through integration into routine quality assurance workflows, ongoing leadership oversight, and continued staff training. Replication in similar low-resource settings is feasible, with necessary adaptation to local context. Future steps include ongoing monitoring, embedding reporting in departmental KPIs, and expanding the project to other hospital units. Further study could explore broader patient-level outcomes, cost-effectiveness, and long-term adherence to reporting practices.

Embedding structured safety reporting within national hospital accreditation frameworks could strengthen patient safety culture across Cambodian hospitals and other LMICs, providing a practical model for replication and continuous improvement.

## Data Availability

All data produced in the present study are available upon reasonable request to the authors

## Abbreviation Full Term

ER: Emergency Room
ICU: Intensive Care Unit
IOB/GYN: Obstetrics and Gynecology
IOPD: Outpatient Department
ILMIC: Low- and Middle-Income Country
ISOP: Standard Operating Procedure
IPDSA: Plan–Do–Study–Act
IBLS: Basic Life Support
IQI: Quality Improvement
IWHO: World Health Organization
INIW: National Interest Waiver (for US immigration context)
ICD: Cesarean Delivery

## Acknowledgments

The author thanks the department heads, nursing leads, and hospital staff for their participation in completing checklists and providing feedback, which supported the implementation and monitoring of this quality improvement initiative. All planning, design, and execution of the project led by the author.

